# *EZH2*: A Critical Competing Endogenous RNA in Cancer Research - A Scoping Review

**DOI:** 10.1101/2024.04.22.24306181

**Authors:** Sadra Salehi-Mazandarani, Sharareh Mahmoudian-Hamedani, Ziba Farajzadegan, Parvaneh Nikpour

## Abstract

In recent years, research on the competing endogenous RNAs (ceRNAs) in cancer is in full swing. These investigations are discovering the importance of critical RNAs in cancer progression. Enhancer of zeste 2 polycomb repressive complex 2 subunit (*EZH2*) is one of these RNAs that has been identified as a potential therapeutic target in many types of cancer. Up to now, many studies have been conducted to elucidate ceRNA role of *EZH2* in cancer. Due to EZH2’s dual role as an oncogene and tumor suppressor in cancer, a more thorough exploration of its ceRNA functions may enhance clinical approaches to cancer treatment. In the current scoping review, we searched online databases including PubMed, Web of Science, Scopus, Embase, Cochrane Library, and Google Scholar to identify experimentally-validated ceRNA axes including *EZH2* in human cancers. We identified 62 unique axes consisting of 30 microRNAs (miRNAs), 31 long non-coding RNAs (lncRNAs), 9 messenger RNAs (mRNAs), and 14 circular RNAs (circRNAs). Notably, *SPRY4-IT1* - miR-101-3p - *EZH2* and *XIST* - miR-101-3p - *EZH2* were recurrent axes observed in multiple cancer types. Among the most frequent miRNAs were miR-101-3p, miR-144-3p, and miR-124-3p, and ceRNAs including *SPRY4-IT1*, *XIST*, *SNHG6*, *HOXA11-AS*, *MALAT1*, and *TUG1* emerged as frequent competitors of *EZH2* for miRNA binding. This scoping review highlights the prevalence and diversity of *EZH2*-containing ceRNA axes in cancer, suggesting their potential as therapeutic targets. Future research should delve deeper into these axes to elucidate their functional significance and assess their clinical applicability.

## 1. Introduction

EZH2 (enhancer of zeste 2 polycomb repressive complex 2 subunit) is the catalytic subunit of the polycomb repressive complex 2 (PRC2), which suppresses gene expression by methylation of lysine 27 of histone 3 (H3K27). This epigenetic modification is thought to silence more than 200 tumor suppressor genes (1). In addition, EZH2 can also methylate non-histone proteins and affect their transcriptional activity. For instance, EZH2 methylates GATA4 transcription factor and reduces its acetylation by p300, resulting in the inhibition of GATA4-mediated transcription (2). Moreover, EZH2 can also activate downstream genes in a PRC2-independent way by methylation of non-histone proteins (3). Through these mechanisms, EZH2 is involved in various cellular processes, such as cell cycle regulation, autophagy, apoptosis, DNA repair and cellular senescence. EZH2 is also associated with many diseases, especially cancer (4).

*EZH2* is overexpressed in many cancers, such as breast, prostate, endometrial and bladder cancers, melanoma, and other malignancies, where it disturbs various cellular processes such as proliferation, apoptosis, migration, and invasion (5). Moreover, EZH2 is subjected to post-translational modifications that modulate its function and activity in cancer. For example, phosphorylation of EZH2 can either promote (pT350-EZH2) or inhibit (pT487-EZH2) cell invasion and metastasis. O-GlcNAcylation, acetylation, and methylation of EZH2 affect proliferation, apoptosis, migration, and metabolism of cancer cells. Ubiquitination of EZH2 can target it for degradation, while deubiquitinases like ZRANB1 stabilize EZH2 and enhance its oncogenic effects (6). However, EZH2 can also act as a tumor suppressor in some cancers, where its loss or inactivation facilitates cancer progression. For instance, T-cell acute lymphoblastic leukemia (T-ALL) is to a large part driven by oncogenic activation of NOTCH1 signaling, which reduces the activity of PRC2 and the histone H3K27me3 repressive mark in T-ALL cells. Inactivation of EZH2 by phosphorylation at its Ser 21 subsequently induces anti-apoptotic genes, such as *IGF1*, *BCL2*, and *HIF1A*, and increases cell adhesion-mediated drug resistance in multiple myeloma cells (7).

*EZH2* expression is also regulated by many microRNAs (miRNAs), which are often deregulated in cancer. Examples of such miRNAs are miR-98 in nasopharyngeal carcinoma (8), miR-101-3p in esophageal squamous-cell carcinoma (ESCC) (9) and miR-124 in gastric cancer (GC) (10). The dysregulation of these miRNAs and their consequent effect on the *EZH2* expression contribute to the aggressive behaviors of these cancers. Therefore, the regulation and role of *EZH2* in cancer is a topic of significant interest to researchers in the field.

RNAs molecules in the cells communicate with each other through shared miRNA response elements (MREs), which act as the building blocks of a novel language and are called competing endogenous RNAs (ceRNAs) (11). Many studies suggest that *EZH2* participates in cancer progression via acting as a member of a ceRNA axis (12–15). As explained by these studies, the RNA of *EZH2*, as part of ceRNA axes, can engage in cancer progression, metastasis, cancer cell proliferation and epithelial-mesenchymal transition (EMT) (Figure 1).

**Figure 1.**
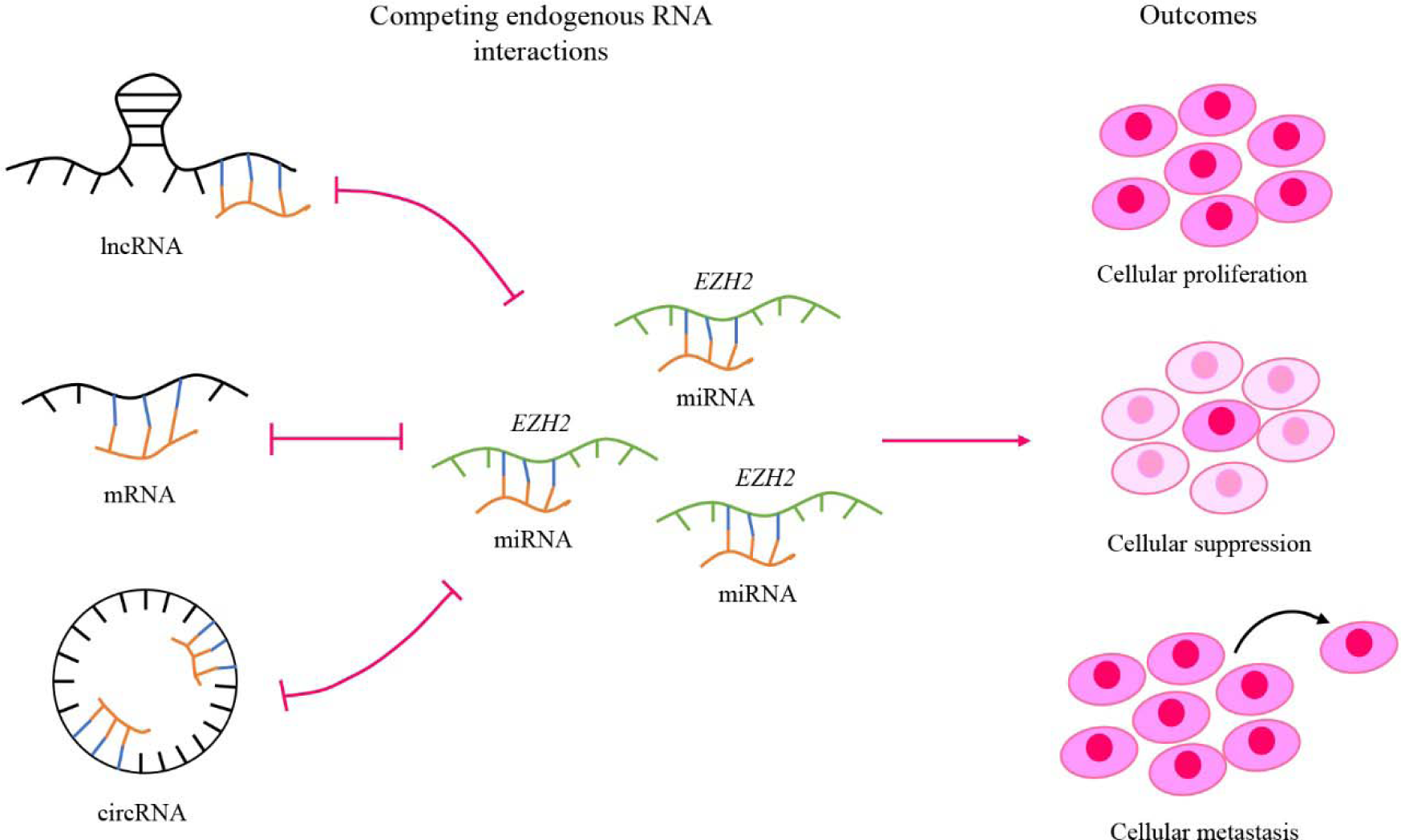
*EZH2*-related competing endogenous RNA (ceRNA) axes and their main outcomes in cancer. *EZH2* competes with ceRNAs for binding to the same miRNAs based on their miRNA response elements (MREs) and forms ceRNA axes. These interactions have critical outcomes in cancer such as affecting on cellular proliferation, suppression, and metastasis.

The current scoping review provides a comprehensive overview of the literature on ceRNA axes including *EZH2* in cancer. While previous studies have exclusively explored the roles of these ceRNA axes in different cancers, this scoping review specifically maps the experimentally validated ceRNA axes containing *EZH2* in cancer and describes the gaps and opportunities regarding employing *EZH2* as a promising targeting component in cancer therapy.

## 2. Materials and methods

This study was performed according to the Preferred Reporting Items for Systematic Reviews and Meta-Analyses statement (PRISMA) (16).

### 2.1. Review Question

Which ceRNA axes containing *EZH2* have been identified in the field of cancer?

### 2.2. Selection criteria

Inclusion criteria included:

1. The *EZH2* RNA has been assessed in any type of cancer.
2. The ceRNA axis has been assessed in human samples.
3. *EZH2* has competition with at least one other ceRNA to sponge a miRNA.
4. The correlations of *EZH2* and other ceRNA(s) with the miRNA(s) have been validated based on the experimental analyses (Not merely bioinformatic-based methods).

Exclusion criteria included:

1. Review studies
2. Studies not in English
3. Retracted studies
4. Studies not specified to cancer
5. Studies in which a specific state of cancer (for example evaluating effects of a drug) have been examined
6. Congresses abstracts

### 2.3. Literature Search Strategy

Online literature search was performed till 6^th^ November 2023 in the PubMed, Web of Science, Scopus, Embase, Cochrane Library, and Google Scholar (first 100 hits) utilizing a developed search query (Supplementary file 1). An attempt was made to contact authors in case of missing information via email.

### 2.4. Screening and data extraction

Records were imported to EndNote software ver. X7 and duplicated studies were removed. Two reviewers (SSM and SMH) independently screened title and abstracts of the studies to select relevant studies. Then, the full texts of the selected studies were assessed against inclusion and exclusion criteria. Consensus was achieved through discussion with another reviewer (PN) to resolve any disagreements.

An excel datasheet was prepared for extracting the data and two reviewers (SSM and SMH) independently performed data extraction from the included studies. Extracted information contained the name of the first author, year of publication, country of origin (the country of the corresponding author), type of cancer, ceRNA axis, method of interaction identification and validation, the main role of the axis, and references. A third reviewer (PN) was consulted in case of disagreement.

### 2.5. Visualization of the ceRNA axes

Cytoscape software (version 3.8.1) (17) was utilized to visualize ceRNA axes including *EZH2* which were identified through the previous steps.

## 3. Results

### 3.1. Characteristics of the included studies

2233 studies were initially identified through online search, among which, 1359 duplicates were omitted. By screening titles and abstracts, 787 studies were excluded, and 27 studies were removed via reading the full texts. Finally, 60 studies were recruited in the current study based on the mentioned criteria. The number of studies at each step of the study selection process is indicated in a PRISMA flowchart (Figure 2). The data extracted from the included studies has been shown in table 1. Excluded studies based on the full text examination with their reason of exclusion are shown in supplementary file 2.

**Figure 2.**
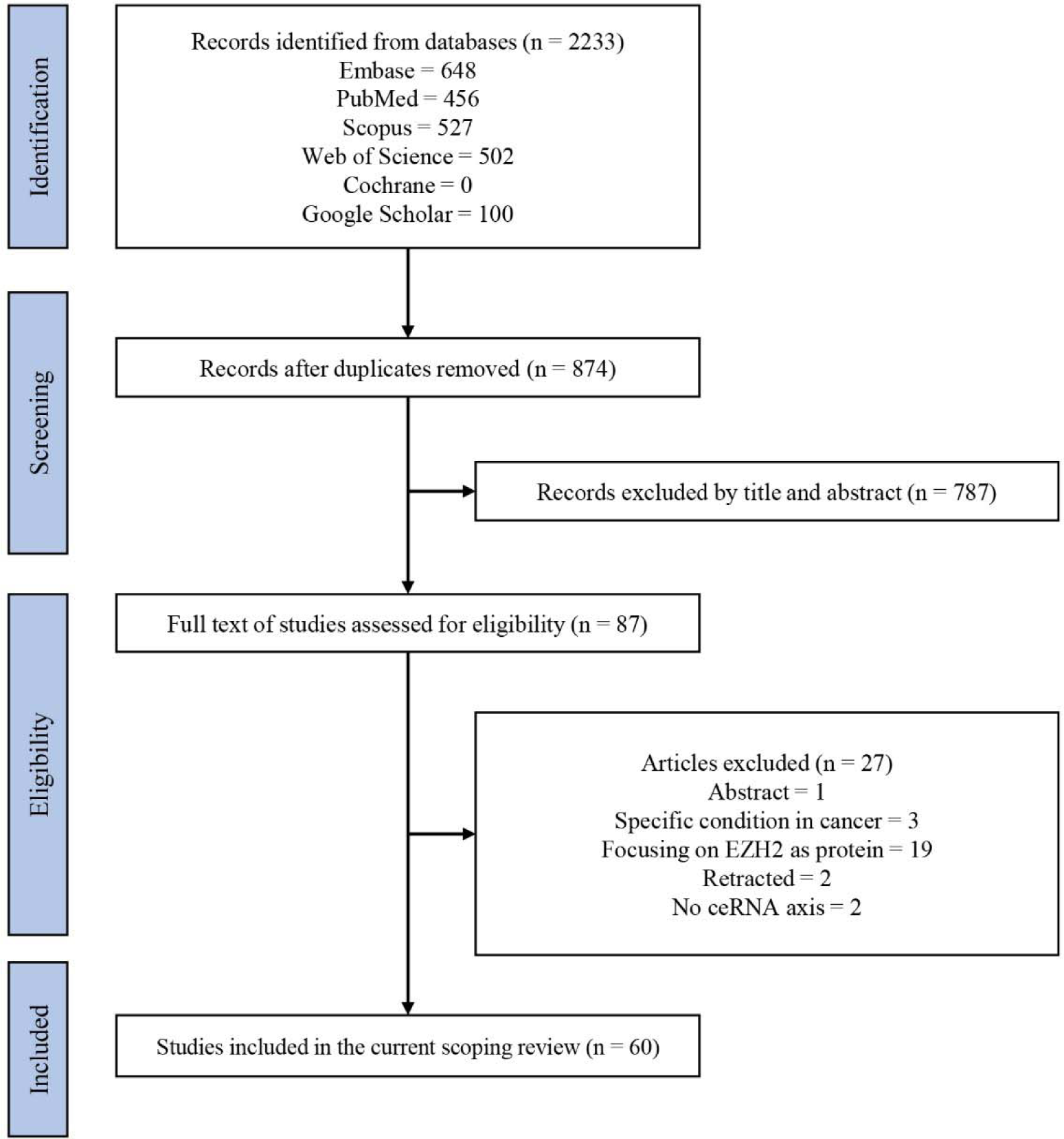
The Preferred Reporting Items for Systematic Reviews and Meta-Analyses (PRISMA) flow diagram of the search results and number of records at each stage, ceRNA: competing endogenous RNA, EZH2: enhancer of zeste 2 polycomb repressive complex 2 subunit

**Table 1.**
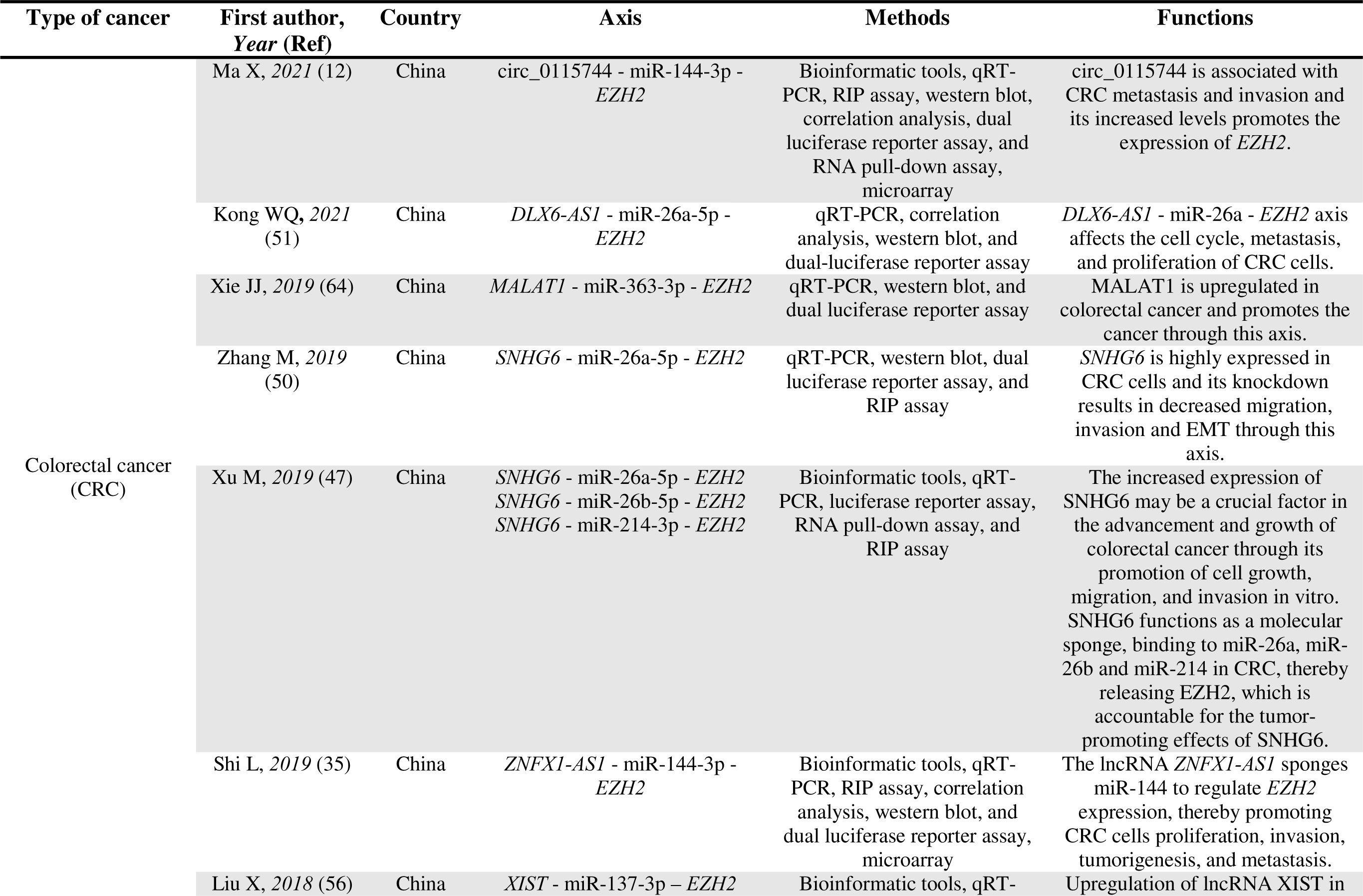
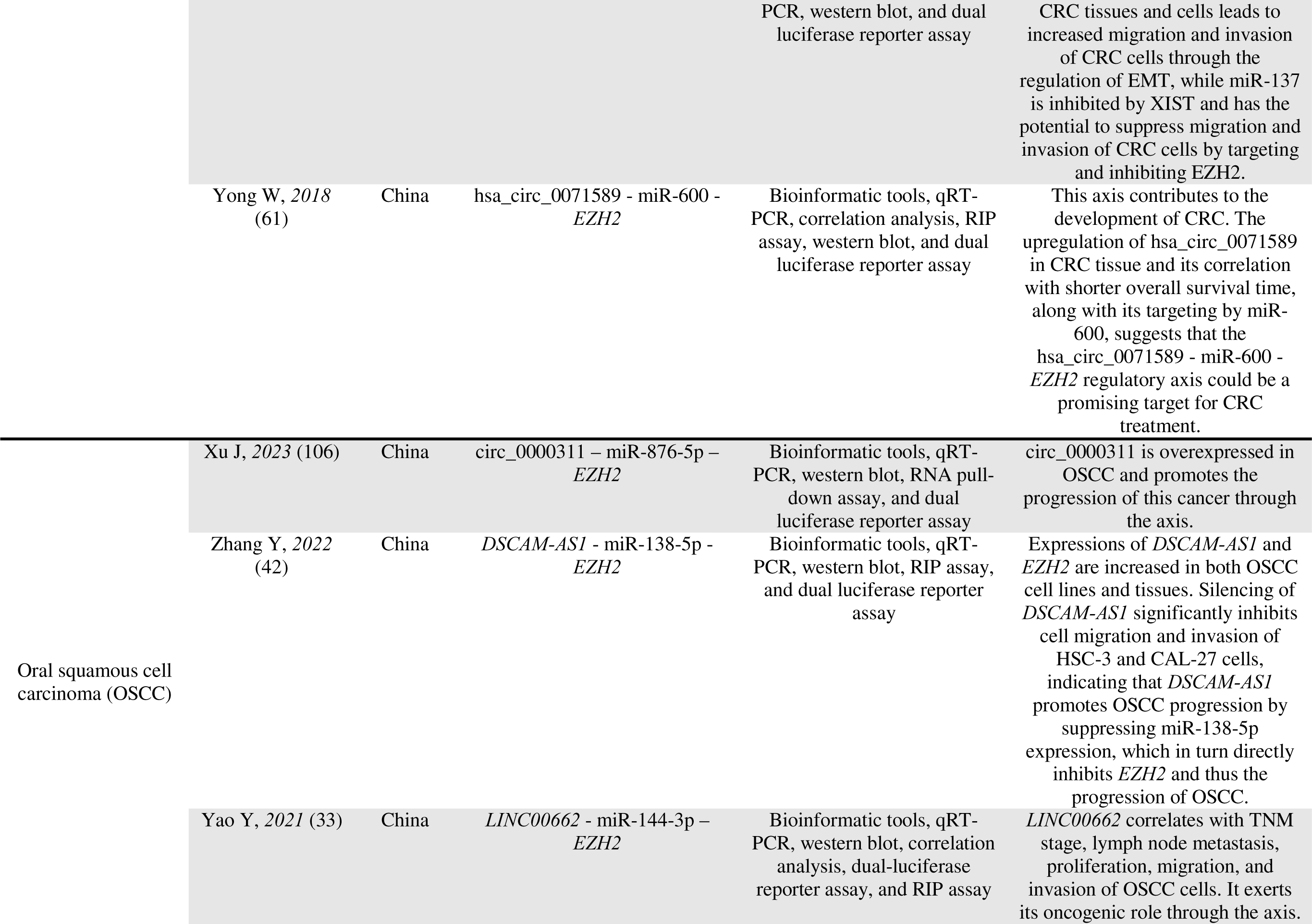
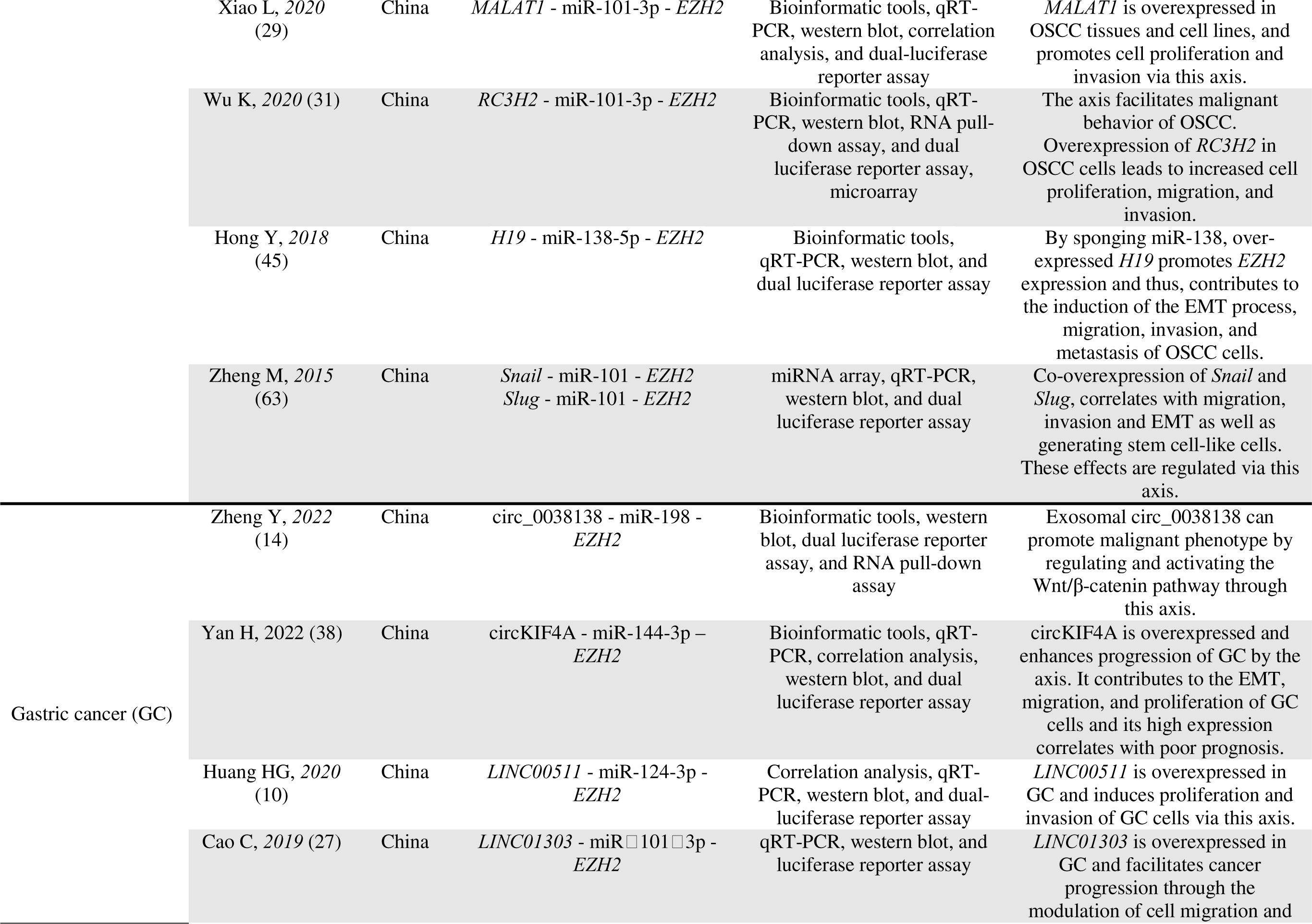
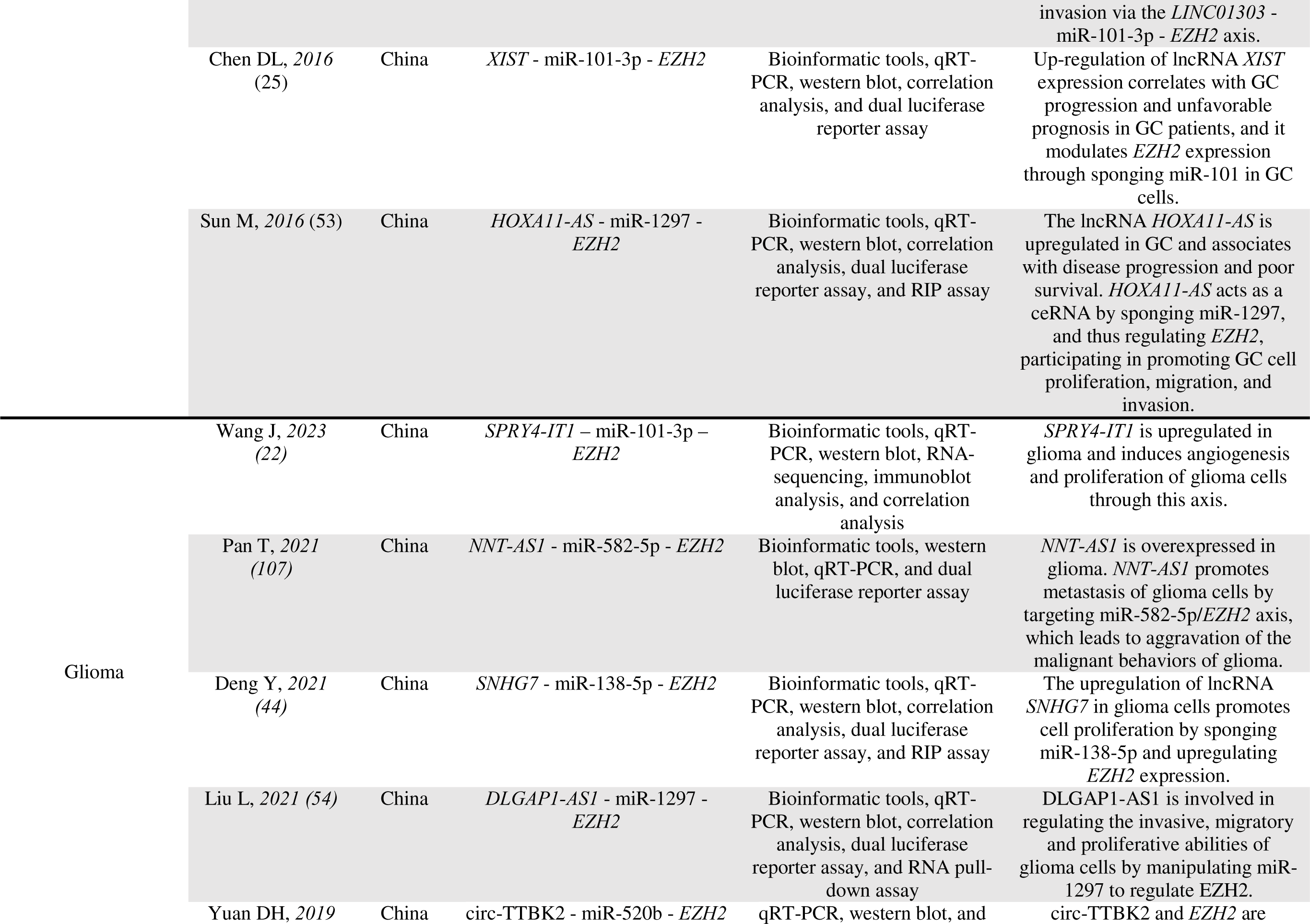
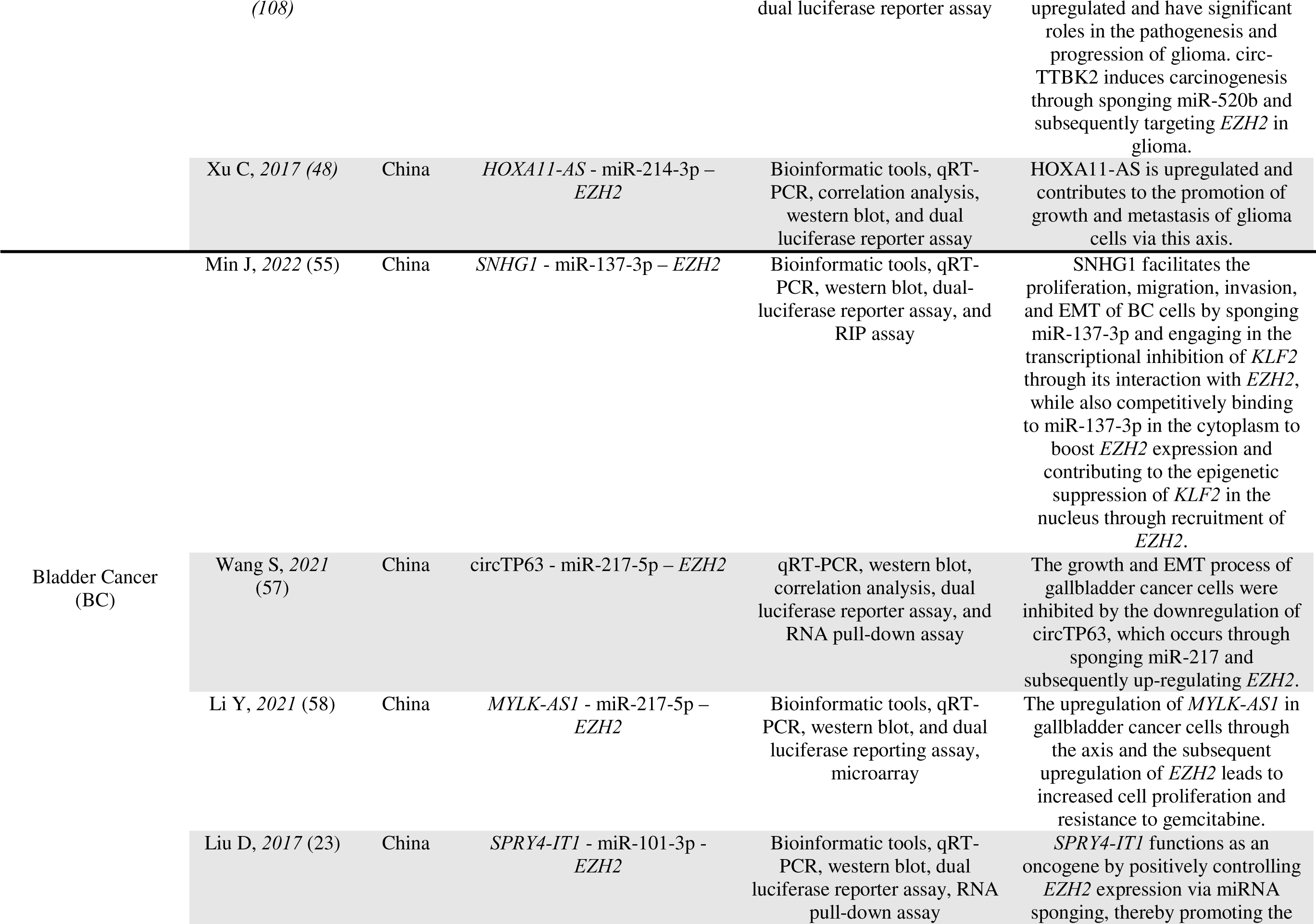
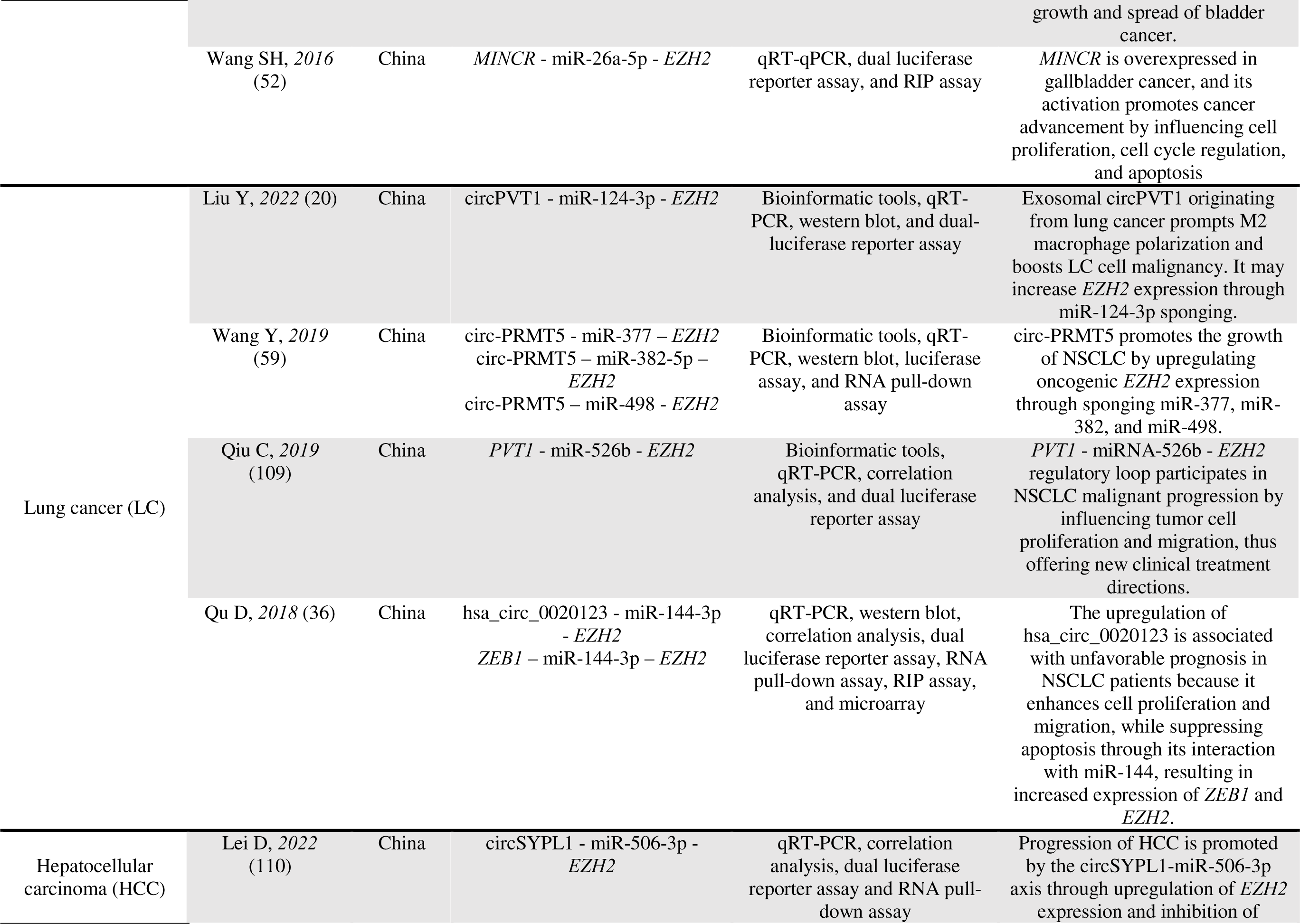
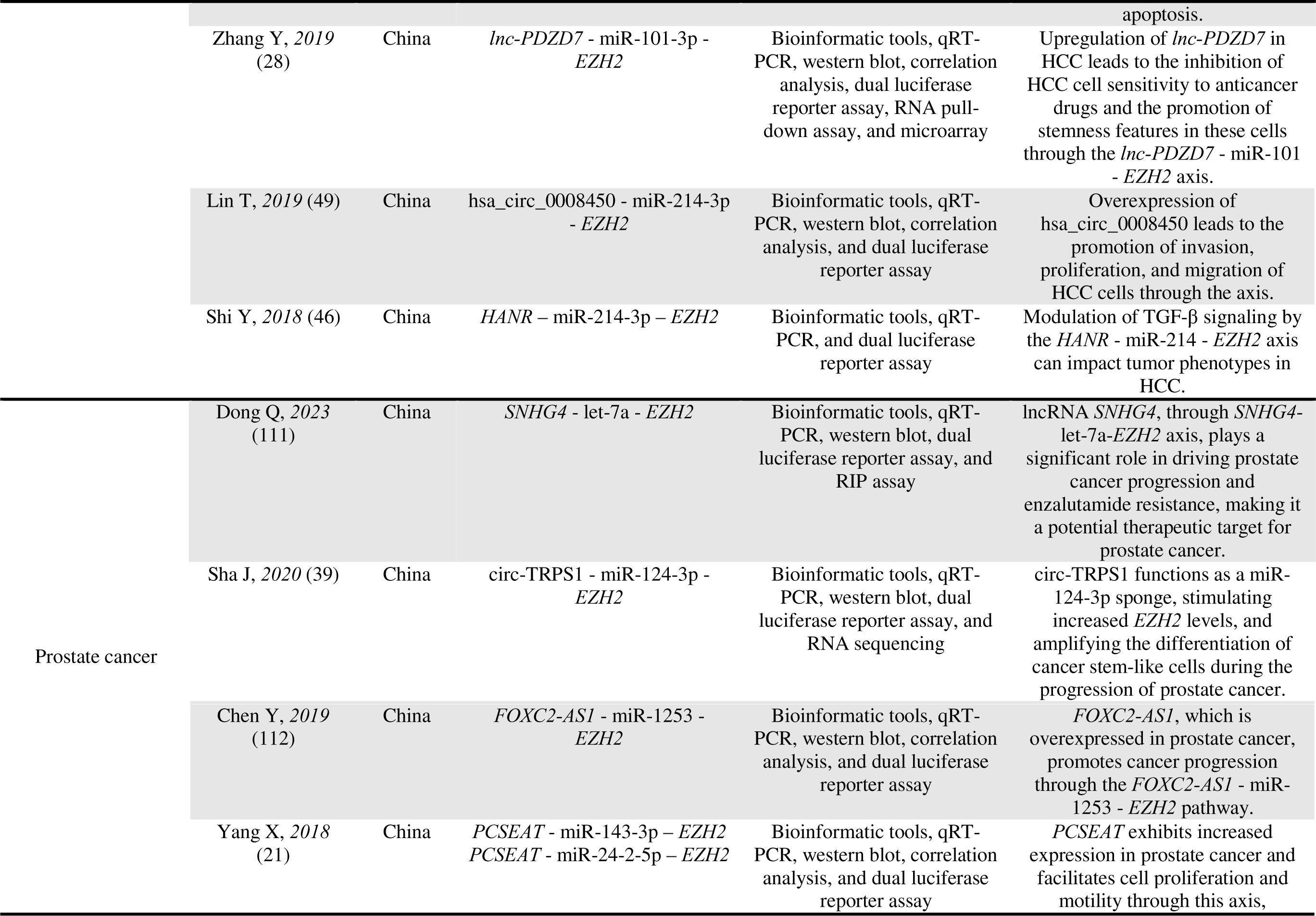
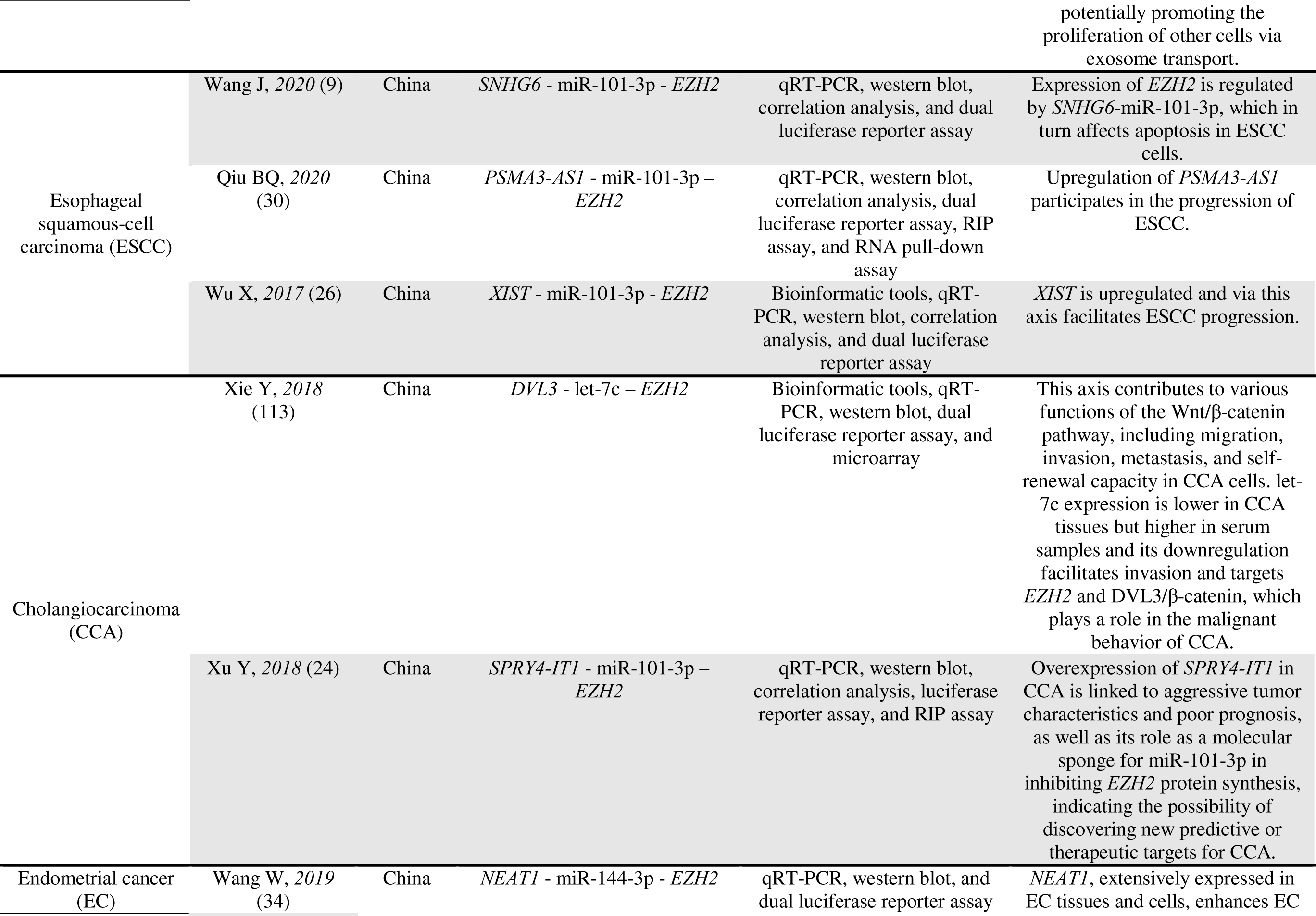
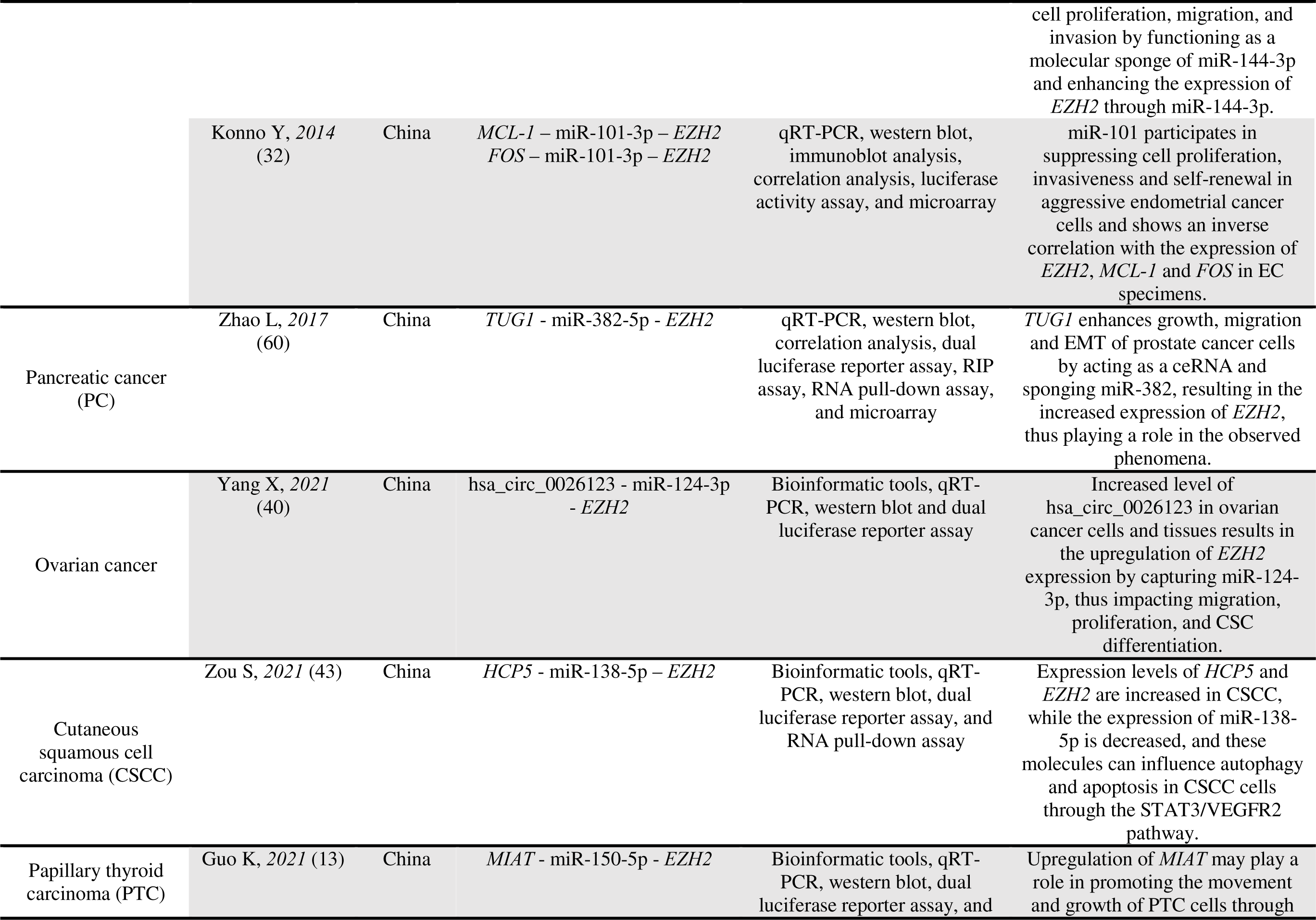
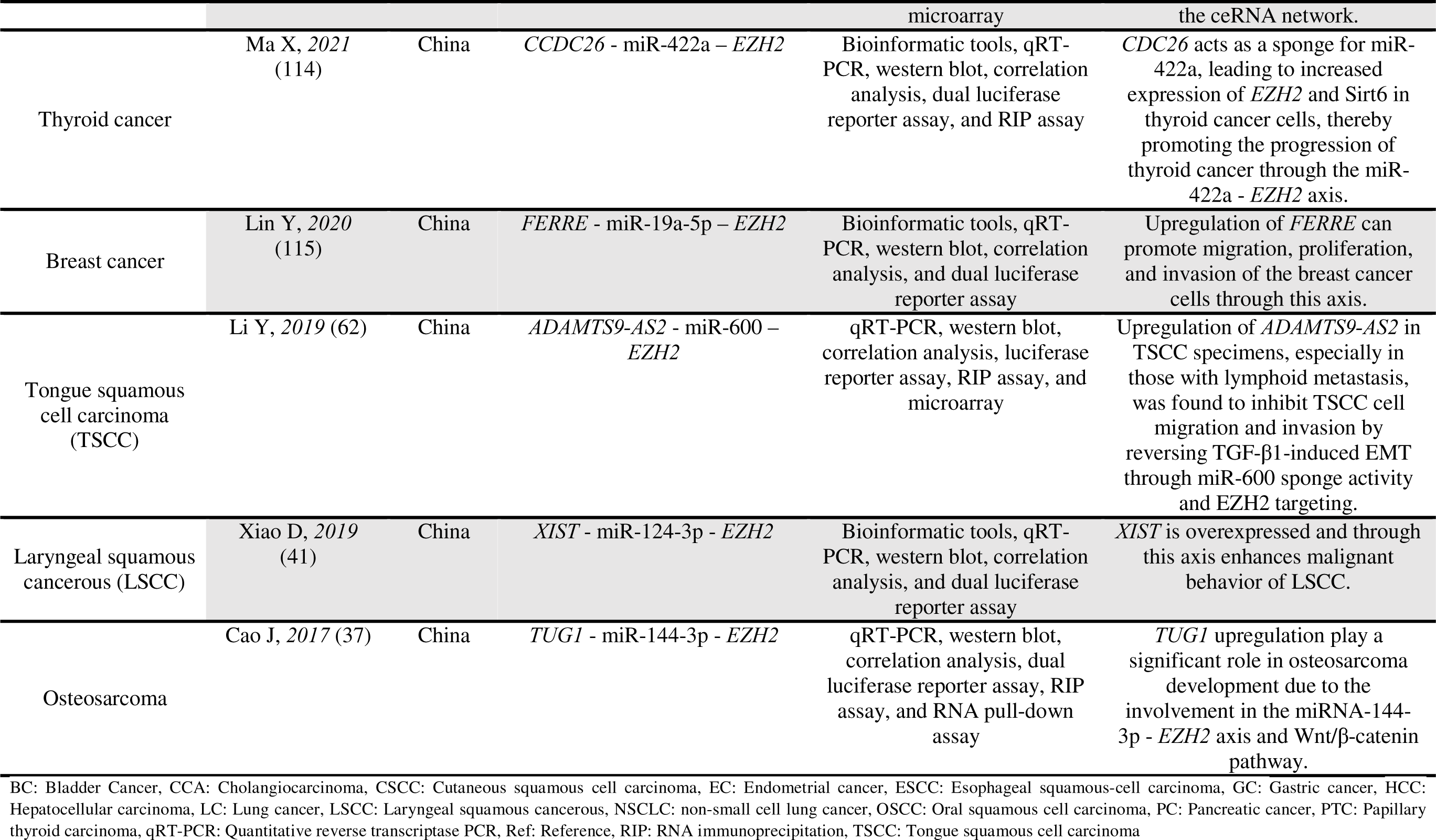
The main characteristics of included studies.

All the included studies were from China and have been conducted between the years of 2014 to 2023. The identified studies have been performed in cancers consist of colorectal, gastric, oral squamous cell carcinoma (OSCC), lung, bladder, glioma, hepatocellular carcinoma (HCC), prostate, esophageal squamous cell carcinoma, pancreatic carcinoma, cholangiocarcinoma, rectal, ovarian, tongue squamous cell carcinoma, cutaneous squamous cell carcinoma, endometrial, papillary thyroid carcinoma, thyroid, laryngeal squamous cancerous, osteosarcoma, and breast. Totally 62 unique axes were identified. There were 30 miRNAs, 31 long non-coding RNAs (lncRNAs), 9 messenger RNAs (mRNAs), and 14 circular RNAs (circRNAs) in the axes. A ceRNA network including these axes has been shown in figure 3.

**Figure 3.**
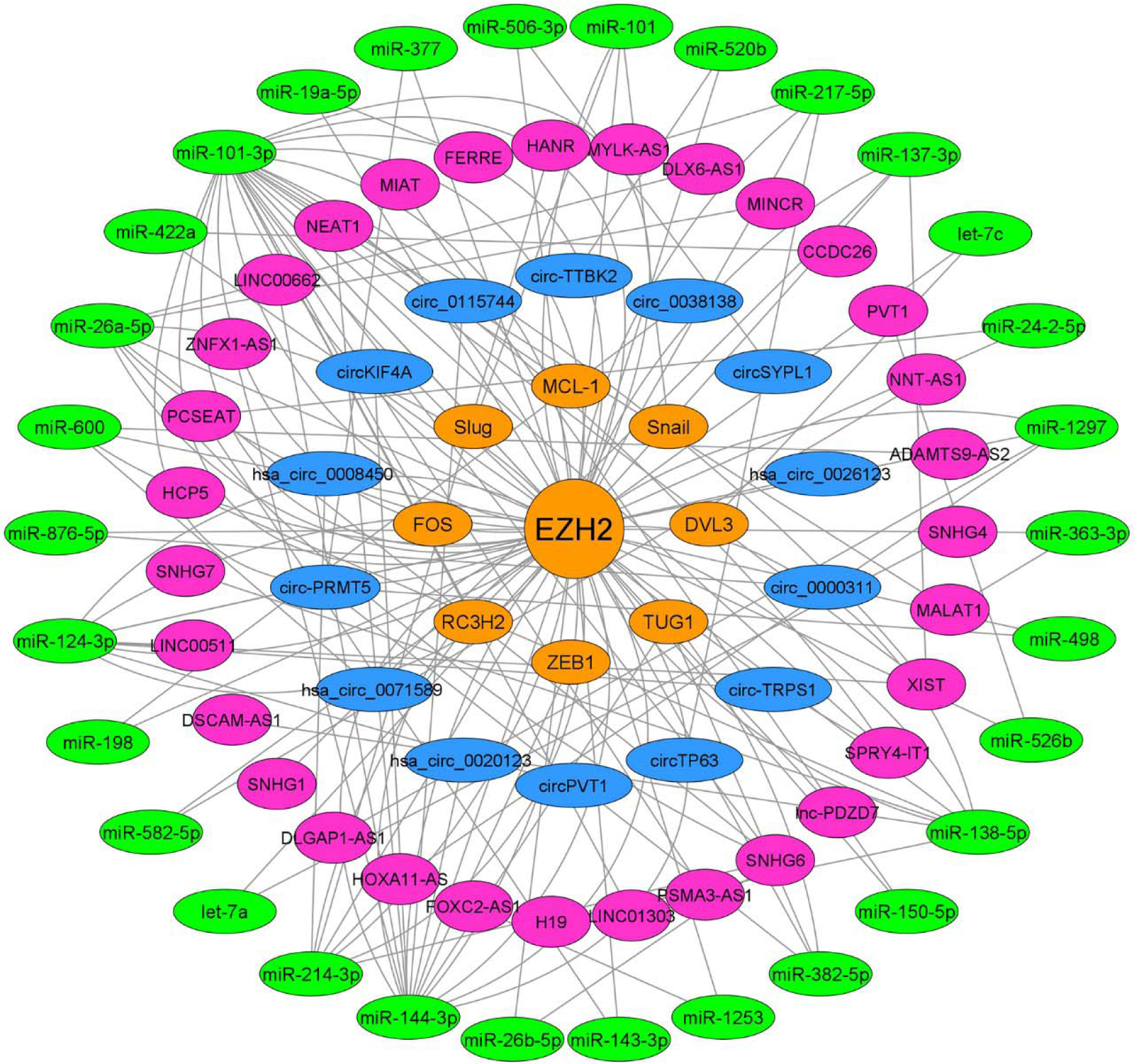
*EZH2*-related ceRNA network in cancer. Orange, blue, purple, and green colors represent mRNAs, circRNAs, lncRNAs, and miRNAs, respectively.

Both experimental and bioinformatic methods have been utilized in the included studies to identify and confirm the interactions among ceRNAs. Experimental methods were such as dual luciferase reporter assay, luciferase reporter, quantitative reverse transcriptase PCR (qRT-PCR), western blot, RNA immunoprecipitation (RIP) and RNA pull-down assays. For bioinformatic analyses, studies used online databases such as TargetScan (18) and CircInteractome (19). In all the included studies, a ceRNA axis including *EZH2*, as a critical ceRNA, as well as the role of this axis on the studied cancer were disclosed. Three of the included studies had examined the ceRNA axes in the context of exosomes (14, 20, 21).

### 3.2. Common *EZH2*-related axes, miRNAs and ceRNAs

*SPRY4-IT1* - miR-101-3p – *EZH2* axis is reported in glioma (22), bladder cancer (23) and cholangiocarcinoma (24). *XIST* – miR-101-3p – *EZH2* was another axis reported in two cancer types including GC (25) and ESCC (26). Eleven miRNAs including miR-101-3p (9, 22–32), miR-144-3p (12, 33–38), miR-124-3p (10, 20, 39–41), miR-138-5p (42–45), miR-214-3p (46–49), miR-26a-5p (47, 50–52), miR-1297 (53, 54), miR-137-3p (55, 56), miR-217-5p (57, 58), miR-382-5p (59, 60), and miR-600 (61, 62) appeared in more than one cancer type/study. Of note, one study (63) did not specify whether they examined the miR-101-3p or other variants of the miR-101 under investigation, so we did not consider it as a common miRNA.

*SPRY4-IT1* (22–24), *XIST* (25, 26, 41, 56), *SNHG6* (9, 47, 50), *HOXA11-AS* (48, 53), *MALAT1* (29, 64), and *TUG1* (37, 60) were ceRNAs repeated in several *EZH2*-related ceRNA axes.

## 4. Discussion

### 4.1. Importance of *EZH2* in cancer

EZH2 is an important regulator in the field of cancer. Many studies have unveiled the roles of EZH2 in different types of cancer. Here we discuss the identified aspects of EZH2 in some selected cancer types.

EZH2 RNA and protein is upregulated in cancerous compared to the non-cancerous colorectal tissues (65). In a meta-analysis in 2017, it was reported that increased expression of EZH2 is associated with better overall survival in patients with colorectal cancer (66). Another study suggested that the inhibition of p21cip1 (also known as P21) expression by EZH2 could result in the activation of the Wnt/β-catenin signaling pathway, which is responsible for regulating the stem-like properties of colorectal cancer cells (67).

EZH2 is overexpressed in cancerous compared to non-cancerous gastric tissues and it correlates with poor prognosis. By regulation of P21 expression, EZH2 affects the proliferation of GC cells (68). Moreover, single nucleotide polymorphisms (SNPs) in *EZH2* correlate with the risk of GC (69, 70). For example, rs734004 genotype CG, rs734005 genotype TC, and rs2072407 genotype TC, all are associated with decreased susceptibility to GC (70). EZH2 promotes cell invasion and progression of GC through activating EMT events and inhibiting cellular senescence (71, 72). Therefore, targeting EZH2 can be a therapeutic method for treating GC (73).

EZH2 is overexpressed in OSCC. Inhibition of EZH2 leads to increase of apoptosis as well as reduction of proliferation, migration and metastasis in this type of cancer (74). Overexpression of EZH2 is associated with shorter survival in patients with OSCC (75). There is also an association between OSCC initiation and the level of EZH2 expression. It has been indicated that high expression of EZH2 in the epithelium of oral leukoplakia, which is the most common potentially malignant disorder of oral mucosa, is linked to a greater risk of developing OSCC (76, 77).

In non-small-cell lung cancer (NSCLC) patients, higher expression of EZH2 correlates with poorer prognosis (78). Expression of EZH2 is also associated with smoking and the expression of KRAS and BRAF in this type of cancer (78). Through suppressing transcription of mesenchymal genes, EZH2 induces mesenchymal to epithelial transition and then tumor colonization in NSCLC (79). The pharmacological values of EZH2 in NSCLC has been reviewed, recently (80).

### 4.2. *EZH2*-related ceRNA axes in cancer

*EZH2* has been identified as a critical ceRNA in various types of cancer. In the current scoping review, we systematically searched and collected the studies in which *EZH2* has been reported as a ceRNA, in a context of axis, by utilizing experimental approaches. Based on the table 1, in some types of cancer, role of *EZH2* as a ceRNA has not been well investigated. However, in some others such as colorectal, gastric and lung cancers, a considerable number of studies have delt with *EZH2* as a critical ceRNA. In these *EZH2*-related ceRNA axes, *EZH2* not only exerts a pivotal influence on the cancer progression, but the ceRNA(s) that compete with *EZH2*, along with the miRNAs present in these axes, play a fundamental role. In fact, the whole axis should be considered as a critical unit. Few of the included studies have been investigated ceRNA axes in exosomes (14, 20). Exosomal ceRNAs are critical for cellular communication in cancer. Their dysregulation affects proliferation, differentiation, and apoptosis of cancerous cells (81).

In the following sections, we discuss common ceRNA axes, miRNAs and ceRNAs among the included studies.

#### 4.2.1. Common ceRNA axes

Among the axes reported in the current study, there were two axes which were common among different cancers. These axes include *SPRY4-IT1* - miR-101-3p – *EZH2* and *XIST* – miR-101-3p – *EZH2*. In this section, we discuss these common axes.

*SPRY4-IT1* - miR-101-3p – *EZH2* ceRNA axis has been reported in glioma, bladder cancer, and cholangiocarcinoma (22–24). *SPRY4-IT1* has been identified as an oncogene in bladder cancer. Its expression correlates with apoptosis, proliferation and migration in this type of cancer (23). *SPRY4-IT1* is overexpressed in cholangiocarcinoma and its expression positively correlates with poor survival. Knockdown of *SPRY4-IT1* in cholangiocarcinoma, promotes apoptosis and decreases proliferation, invasion and migration (24). In these cancers, *SPRY4-IT1* regulates the expression of *EZH2* via sponging miR-101-3p (22–24).

*XIST* – miR-101-3p – *EZH2* ceRNA axis has been reported as a critical axis in GC and esophageal squamous cell carcinoma with *XIST* upregulation reported in both cancers (25, 26). In GC, high expression of *XIST* positively correlates with tumor size, tumor, node, metastasis (TNM) stage, invasion, and metastasis (25). In esophageal squamous cell carcinoma, *XIST* expression associates with invasion, proliferation, and migration of cancerous cells and patients with its higher expression, have shorter survival (26). In both cancers, *XIST* expression can be regulated by *EZH2* through sponging miR-101-3p (25, 26).

The recurrence of these two commonly observed ceRNA axes across various types of cancer emphasizes their crucial role in the development of cancer. These axes serve as intricate regulatory networks, orchestrating gene expression patterns that drive tumorigenesis and progression. The fact that they are found in different cancer types suggests a consistent mechanism of disruption in cancer cells, which presents promising opportunities for targeted interventions. Utilizing these ceRNA axes as diagnostic biomarkers or therapeutic targets has great potential for enhancing precision oncology strategies. Further investigation into their mechanistic underpinnings and clinical implications is warranted to harness their full therapeutic utility in combating cancer.

#### 4.2.2. Common miRNAs in the axes

Among the included studies, there were common miRNAs interacting with *EZH2* in various types of cancer or in one type based on different studies. Eleven miRNAs including miR-101-3p, miR-144-3p, miR-124-3p, miR-138-5p, miR-214-3p, miR-26a-5p, miR-1297, miR-137-3p, miR-217-5p, miR-382-5p, and miR-600 were the common miRNAs. Here we briefly discuss the implication of these miRNAs in different cancer types.

In twelve included studies, which were conducted on OSCC (29, 31), esophageal squamous cell carcinoma (9, 26, 30), GC (25, 27), HCC (28), bladder cancer (23), endometrial cancer (32), cholangiocarcinoma (24), and glioma (22), miR-101-3p has shown a correlation with *EZH2*. miR-101-3p has been reported as a critical miRNA in cancer. miR-101-3p acts as a tumor suppressor in NSCLC through targeting *MALAT-1* and PI3K/AKT signaling pathway (82). In GC, miR-101-3p exhibits tumor suppressor properties by inhibiting cell proliferation and induction of apoptosis via regulation of *PIM-1* expression (83). However, there are also some studies suggesting that miR-101 functions as an oncogene, particularly in vivo, and increased levels of miR-101-3p have been associated with poorer survival outcomes in ovarian cancer patients (84). Detailed cancer-related crucial roles of miR-101-3p has been reviewed elsewhere (85).

Seven of the included studies reported ceRNA axes including miR-144-3p. These studies were performed in cancers including colorectal (12, 35), lung (36), gastric (38), endometrial (34), OSCC (33), and osteosarcoma (37). miR-144-3p is dysregulated in cancers and can have both tumor suppressive and oncogenic roles depending on its target genes (86).

miR-124-3p has been reported in five of the included studies. These studies have been conducted in cancers including gastric (10), lung (20), prostate (39), ovarian (40), laryngeal squamous cell carcinoma (41). miR-124-3p has association with initiation, progression, and survival in cancer (87).

miR-138-5p and miR-214-3p have been reported, separately, in four studies. miR-138-5p have been reported in OSCC (42, 45), glioma (44) and cutaneous squamous cell carcinoma (43). miR-214-3p have been presented in HCC (46, 49), glioma (48), and colorectal cancers (47). miR-138-5p has been reported as a tumor suppressive miRNA preventing proliferation of cancerous cells. It performs this role by binding to the mRNAs producing oncogenic proteins (88). miR-214-3p is dysregulated in cancer and can both induce or prevent the proliferation of cancerous cells (89).

miR-26a-5p has been reported in four studies. The studies were performed in colorectal (47, 50, 51) and bladder cancers (52). The importance of this miRNA has been reported in different cancers. For instance, circulating levels of miR-26a-5p in plasma of patients with pediatric rhabdomyosarcoma are downregulated. Furthermore, patients with higher expression of miR-26a-5p have longer overall and progression free survival in this type of cancer (90). miR-26a-5p is also downregulated in breast cancer tissues. It can inhibit proliferation, sensitivity to chemotherapy, and metastasis of breast cancer cells (91).

miRNAs including miR-1297 (53, 54), miR-137-3p (55, 56), miR-217-5p (57, 58), miR-382-5p (59, 60), and miR-600 (61, 62) separately, were reported in two studies. The important association of these miRNAs with cancer progression have been uncovered (92–96). For example, miR-1297 expression is deregulated in cancer and affect tumorigenesis by playing oncogenic or tumor-suppressive roles (92). miR-217-5p is also dysregulated in many types of cancer and its expression is influenced by various upstream regulators including lncRNAs and circRNs (93).

#### 4.2.3. Common ceRNAs in the axes

Among the ceRNAs in the axes extracted from included studies, some were more frequent than others, including *SPRY4-IT1, XIST*, *SNHG6*, *HOXA11-AS*, *MALAT1*, and *TUG1* which we briefly discuss these RNAs in this section.

*SPRY4-IT1* has been reported in three of the included studies (22–24) and in all of them it competes with *EZH2* for binding to the miR-101-3p. lncRNA *SPRY4-IT1* is a critical RNA in cancer. For instance, it is overexpressed in prostate cancer and through the PI3K/AKT signaling pathway, promotes proliferation of cancerous cells (97). In breast cancer, upregulation of *SPRY4-IT1* induces NF-κB signaling pathway by which promotes cell proliferation and inhibits apoptosis (98).

*XIST* has been reported in four studies. It is reported to compete with *EZH2* for binding to miR-101-3p in GC (25) and ESCC (26) miR-137-3p in colorectal cancer (56), and miR-124-3p in laryngeal squamous cell carcinoma (41). The alteration of *XIST* expression in cancer has significant implications for cell growth, invasion, response to chemotherapy, and metastasis. The mechanisms by which it exerts these roles have been reviewed elsewhere (99, 100).

*SNHG6* has been reported in three of the included studies. These studies show that *SNHG6* competes with *EZH2* for binding to the miR-214-3p and miR-26a/b in colorectal cancer (47, 50) and miR-101-3p in esophageal squamous cell carcinoma (9). A meta-analysis, published in 2020, has reported the correlation of *SNHG6* expression with advanced TNM stage, tumor invasion depth, lymph node metastasis, and distance metastasis in human cancers (101).

*HOXA11-AS* has been reported as a *EZH2* competitor in GC (53) and glioma (48). *HOXA11-AS* participates in cancer development through various mechanisms such as epigenetic modifications in the nucleus and acting as a ceRNA in the cytoplasm (102) which have made it a promising therapeutic target in the field of cancer.

*MALAT1* is another lncRNA which is reported as a *EZH2* competitor in OSCC (29) and colorectal cancer (64). *MALAT1* is mostly overexpressed in human cancers and its expression is associated with tumor size and stage as well as patients’ poor survival (103). *MALAT1* has been recently identified as an important RNA which enhances cellular resistance to chemotherapy (104).

Two of the included studies have reported *TUG1* as a competitor of *EZH2* to bind to miR-144-3p and miR-382-5p in osteosarcoma (37) and pancreatic cancer (60), respectively. *TUG1* exhibits abnormal expression in human cancers and, by recruiting specific RNA-binding proteins, it induces the expression of target genes. Additionally, it functions as an important ceRNA, affecting the development of cancerous cells (105).

### 4.3. Strengths, limitations, and suggestions for the future studies

The current scoping review study possesses several strengths. First, different databases were systematically searched. Furthermore, we focused on experimentally validated ceRNA axes rather than those only predicted through prediction algorithms. Finally, we searched ceRNA axes in all types of cancers and provided a comprehensive view of *EZH2* as a ceRNA in cancer.

As for the limitation of the current scoping review, there are various constraints that need to be considered when interpreting the findings, including the absence of relevant articles in certain types of cancers like melanoma and kidney cancer, as well as the potential publication bias regarding studies with null or negative findings.

This scoping review presents recommendations for future research. An imperative exists for the thorough exploration of common ceRNA axes, miRNAs and ceRNAs, given their considerable significance in cancer biology. It is essential to delve deeper into their underlying mechanisms to fully exploit their therapeutic potential in the management of cancer. On the other hand, while *EZH2* has been identified as a crucial ceRNA in certain cancers, its role remains insufficiently studied in others, thus necessitating further investigation to gain insights into the molecular characteristics of EZH2 in other cancer types. Furthermore, exploring these axes in exosomes could provide valuable indications of their relevance as a prominent subject in cancer research.

## 5. Conclusions

Multiple lines of evidence have reported vital functions of *EZH2* in cancers. In this scoping review, we summarized the emerging insights on ceRNA axes including *EZH2* in cancer which have been experimentally validated. These axes contain competition of *EZH2* with other critical ceRNAs for binding to the miRNAs, thereby influencing cancer progression. The current study provides a comprehensive perspective of these ceRNA axes in cancer focusing on *EZH2*. Further studies are needed to further investigate these axes and assess their clinical utilities.

## Data Availability

All data produced in the present study are available upon reasonable request to the authors.

## Author contributions

**Sadra Salehi-Mazandarani:** Conceptualization, Methodology, Investigation, Writing - original draft. **Sharareh Mahmoudian-Hamedani:** Methodology, Investigation, Writing - original draft. **Ziba Farajzadegan:** Writing - review and editing, Supervision, Validation. **Parvaneh Nikpour:** Funding acquisition, Supervision, Project administration, Writing - review and editing, Validation.

