## Supplementary material for "*EZH2*: A Critical Competing Endogenous RNA in Cancer Research - A Scoping Review": Suppl. File 1

| Database | Search query | Number of identified records until  6 Nov 2023 |
| --- | --- | --- |
| PubMed, all fields | (cerna* OR "competing endogenous RNA*" OR "competitive endogenous RNA*" OR "microRNA response element*" OR "miRNA response element*" OR sponge OR axis) AND (EZH2 OR "Enhancer of Zeste Homologue 2") AND (cancer OR malignancy OR neoplasm* OR tumor OR tumour) | 456 |
| Web of Science, topic | TS=((cerna* OR "competing endogenous RNA*" OR "competitive endogenous RNA*" OR "microRNA response element*" OR "miRNA response element*" OR sponge OR axis) AND (EZH2 OR "Enhancer of Zeste Homologue 2") AND (cancer OR malignancy OR neoplasm* OR tumor OR tumour)) | 502 |
| Scopus, title abstract keyword | TITLE-ABS-KEY((cerna* OR "competing endogenous RNA*" OR "competitive endogenous RNA*" OR "microRNA response element*" OR "miRNA response element*" OR sponge OR axis) AND (EZH2 OR "Enhancer of Zeste Homologue 2") AND (cancer OR malignancy OR neoplasm* OR tumor OR tumour)) | 527 |
| Embase, all fields | (cerna* OR "competing endogenous RNA*" OR "competitive endogenous RNA*" OR "microRNA response element*" OR "miRNA response element*" OR sponge OR axis) AND (EZH2 OR "Enhancer of Zeste Homologue 2") AND (cancer OR malignancy OR neoplasm* OR tumor OR tumour) | 648 |
| Cochrane Library, title abstract | ((cerna* OR "competing endogenous RNA*" OR "competitive endogenous RNA*" OR "microRNA response element*" OR "miRNA response element*" OR sponge OR axis) AND (EZH2 OR "Enhancer of Zeste Homologue 2") AND (cancer OR malignancy OR neoplasm* OR tumor OR tumour)):ti,ab,kw | 0 |
| Google Scholar | ceRNA axis including EZH2 in cancer | 100^*^ |

^*^ First 100 records
