## Supplementary material for "*EZH2*: A Critical Competing Endogenous RNA in Cancer Research - A Scoping Review": Suppl. File 2

**Excluded studies based on full text examination**

| **Reason of exclusion** | **References** |
| --- | --- |
| Abstract | (1) |
| A specific condition in cancer | (2-4) |
| Focusing on the EZH2 as a protein (epigenetic regulator) | (5-23) |
| Retracted | (24, 25) |
| No ceRNA axis | (26, 27) |
